# Global Monitoring of the Impact of COVID-19 Pandemic through Online Surveys Sampled from the Facebook User Base

**DOI:** 10.1101/2021.07.05.21259989

**Authors:** Christina M. Astley, Gaurav Tuli, Kimberly A. Mc Cord - De Iaco, Emily L. Cohn, Benjamin Rader, Samantha Chiu, Xiaoyi Deng, Kathleen Stewart, Tamer H. Farag, Kris M. Barkume, Sarah LaRocca, Katherine A. Morris, Frauke Kreuter, John S. Brownstein

**Author notes:** Corresponding author Christina M. Astley, MD, ScD, **Email:**. These authors contributed equally (Christina M. Astley, Gaurav Tuli, Kimberly A. Mc Cord - De Iaco). These authors contributed equally (Frauke Kreuter, John S. Brownstein). **Author Contributions:** CMA, JSB, and FK designed the research. CMA, KA Mc Cord - De Iaco, BR, and GT performed research. KMB, SC, XD, THF, FK, SLR, KA Morris contributed new data and analytic tools. CMA, KA Mc Cord - De Iaco, BR, and GT analyzed data. CMA, KAM (Mc Cord - De Iaco) drafted the manuscript. All authors revised and approved of the paper. **Competing Interest Statement:** Nothing to disclose.

## Abstract

Simultaneously tracking the global COVID-19 impact across multiple populations is challenging due to regional variation in resources and reporting. Leveraging self-reported survey outcomes via an existing international social media network has the potential to provide reliable and standardized data streams to support monitoring and decision-making world-wide, in real time, and with limited local resources. The University of Maryland Global COVID Trends and Impact Survey (UMD-CTIS), in partnership with Facebook, invites daily cross-sectional samples from the social media platform’s active users to participate in the survey since launch April 23, 2020. COVID-19 indicators through December 20, 2020, from N=31,142,582 responses representing N=114 countries, weighted for nonresponse and adjusted to basic demographics, were benchmarked with government data. COVID-19-related signals showed similar concordance with reported benchmark case and test positivity. Bonferroni significance and minimal Spearman correlation strength thresholds were met in the majority. Light Gradient Boost machine learning trained on national and pooled global data verified known symptom indicators, and predicted COVID-19 trends similar to other signals. Risk mitigation behavior trends are correlated with, but sometimes lag, risk perception trends. In regions with strained health infrastructure, but active social media users, we show it is possible to define suitable COVID-19 impact trajectories. This syndromic surveillance public health tool is the largest global health survey to date, and, with brief participant engagement, can provide meaningful, timely insights into the COVID-19 pandemic and response in regions under-represented in epidemiological analyses.

**Significance Statement:** The University of Maryland Global COVID Trends and Impact Survey (UMD-CTIS), launched April 23, 2020, is the largest remote global health monitoring system. This study includes about 30 million UMD-CTIS responses over 34 weeks (through December 2020) from N=114 countries with survey-weights to adjust for nonresponse and demographics. Using limited self-reported data, sampled daily from an international cohort of Facebook users, we demonstrate validity and utility for COVID-19 impacts trends, even in regions with scant or delayed government data. We predict COVID-19 cases in the absence of testing, and characterize perceived COVID-19 risk versus risk-lowering measures. The UMD-CTIS has the potential to support existing monitoring systems for the COVID-19 pandemic, as well as other new, as-yet-undefined global health threats.

## Introduction

In December 2019, the COVID-19 pandemic swept across the globe and challenged the scientific community to urgently assess and intervene(1). While impressive public health mitigation efforts have been made, such as through expansion of case testing and reporting infrastructure, intensive non-pharmaceutical interventions and rapid vaccine development, barriers remain to understanding and controlling the pandemic (2). The lack of pre-existing knowledge of the SARS-CoV-2 virus and lack of uniform COVID-19 data among and within countries has reduced our ability to assess severity and direct action across regions(3). This data issue may have been doubly challenging in areas with less resilient healthcare systems, where disease surveillance may be delayed or underestimated and surge capacity limited(4, 5). This lack of timely information may obscure the actual impact during a crucial window to mobilize targeted support(6).

One successful approach has been to leverage syndromic surveillance through digital surveys(7–12). Previous work on influenza and now COVID-19 has demonstrated utility of this approach based on self-reported symptoms and other data, in near real-time, including for burden estimation, hot-spots identification, and mitigation tracking(13–16). While platforms differ in survey methodology, scope and source population, insights have been comparable and used to inform decision-making, thus further underscoring the value of participatory epidemiology in the setting of pandemic.

While existing platforms have many strengths, such as longitudinal disease trajectories(9, 17), rapid deployment of timely survey questions(11, 18), there is a need for sufficient data from diverse populations, especially from regions already struggling with pandemic response. High-quality, prospective, opt-in platforms that rely on government endorsement, advertising, or word-of-mouth may not be feasible with low-uptake, disease incidence or long-term engagement, and may be particularly problematic in populations with limited government trust or variable access to smartphone technology.

Towards this end, the University of Maryland (UMD) and Facebook developed the COVID-19 Trends and Impact Survey (UMD-CTIS), a lower-density sampling scheme deployed using a novel recruitment mechanism to a massive social media platform source population — the Facebook Active User Base (FAUB) — that can be mapped back to the national population for 114 countries worldwide(19). Thus, because the FAUB spans countries with varied languages, social structures and economic resources, UMD-CTIS provides a unique understanding of global and region-specific health at a scale not feasible previously(20). Utilizing data from the era prior to COVID-19 vaccine deployment, we benchmark epidemiologic parameters, use human social sensing signals(21) and predicted SARS-CoV-2 test positivity for regions with limited testing. We also illustrate perception of pandemic risk-mitigation trade-offs trends in under-represented regions.

## Results

### Study Participants and Response Characteristics

The UMD-CTIS (summarized in **Figure 1**) characterized 127 self-reported time-varying COVID-19-related measures including key demographic, behavior and health impacts at an unparalleled spatiotemporal scale. Consistent survey methodology was applied through 6 versions and across 250 countries via a unified instrument in 56 languages. The survey was available daily from April 23 to December 20, 2020. In this analysis of the first 6 versions, we include N=31,142,582 cross-sectional surveys completed over the course of 34 ISO weeks (April 27 to December 20, 2020), with a median [interquartile range (IQR)] of 7,837 [6,312, 9,458] surveys per week per country for the N=114 countries with survey weights. Surveys required limited time commitment with a median [IQR] duration of 9.2 [5.3,10.1] minutes. Sample size (log10, black), coverage (percent surveys per population, white), age and gender distributions by country are shown as bar charts in **Figure 2** (survey-weighted in **Supplementary Figure 1**). Male respondents are more frequent in Africa and Asia, in contrast to the documented female-predominance of this and other European health research studies, and may reflect regional differences in technology access and use (22, 23). Survey representativeness compared to the COVID-19-related biomedical literature is higher in Central and South America, Eastern Europe and parts of Asia (**Figure 1**).

**Figure 1.**
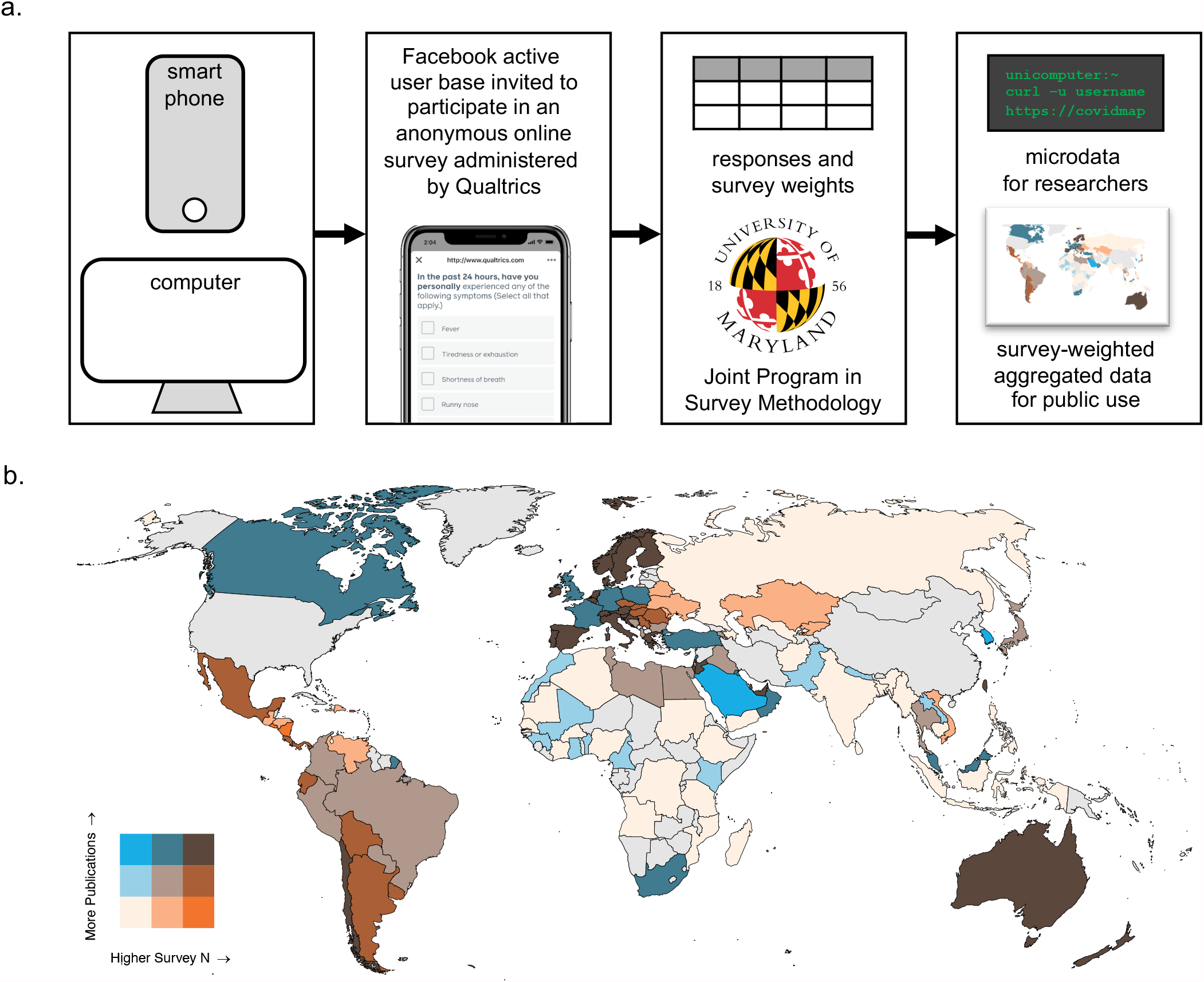
Data acquisition pipeline and survey representativeness overview. a) The Facebook Active User Base is sampled daily, and invited to participate in the online UMD-CTIS, administered by Qualtrics, accessed via an online form using a smartphone or computer. Participants are asked about demographics, COVID-19 symptoms, behaviors, and outcomes. Facebook supplies survey weights to account for non-response and to adjust for basic demographics of the participant. Aggregated data are released to the public in near-real-time. Researchers may apply to use raw microdata to study COVID-19. b) Representativeness of the UMD-CTIS survey in each country relative to representation in the COVID-19 biomedical literature. UMD-CTIS surveys per capita (orange) versus COVID-19-related publications per capita (blue) for the N=114 countries with survey-weights in this analysis.

**Figure 2.**
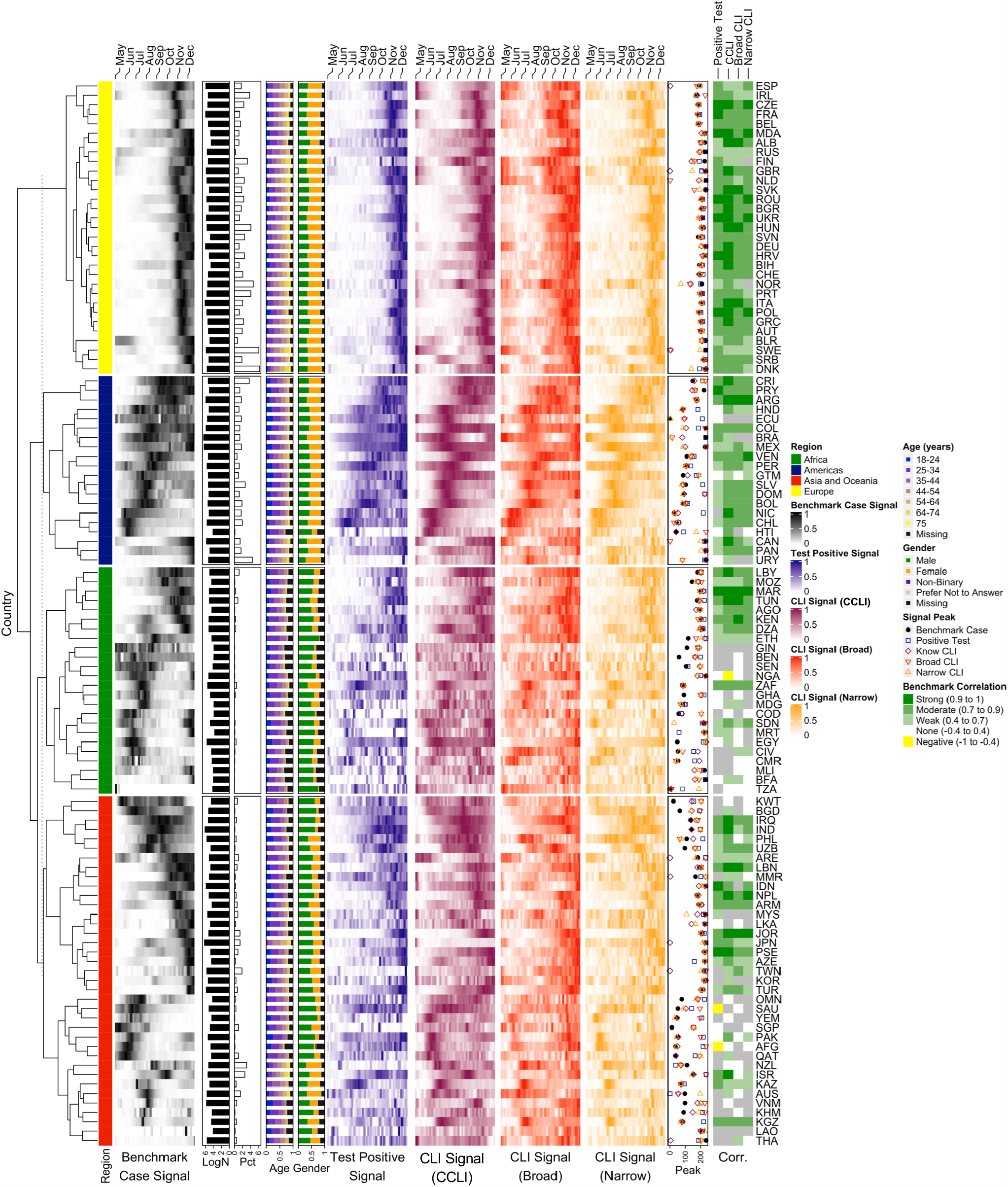
Time series heatmap comparing the benchmark signals to UMD-CTIS-based signals. Within-country seven-day smoothed benchmark and signal data are normalized to range [0,1] using [minimum, maximum]. Clustering within regions using benchmark data. Demographics: log10 surveys (*LogN*) and proportion of surveys per population (*pct*) as bar charts, and proportion of surveys in for each age (*Age*) or gender (*Gender*) as stacked bar charts. Signals shown include recent positive COVID-19 (*Test Positive*), community CLI (CCLI), self-reported fever, cough or anosmia/ageusia (*Broad CLI*), or self-reported anosmia/ageusia of less than 14 days duration (*Narrow CLI*). The time-to-first-peak in days for each signal is compared (*Peak*). Spearman correlation of benchmark cases with each UMD-CTIS signal summarized by strength of the correlation for those with p-values surviving Bonforroni correction (e.g. < 0.05/448).

### Benchmarking Trends in COVID-19 Impact

The unique methodology of this survey, leveraging the global FAUB and national census demographics, allows for estimation of daily population-standardized health metric trajectories for N=114 countries with relatively few, generally brief, one-time, cross-sectional surveys(19). As a proof of principle (**Figure 2**), we sought to test whether benchmark Oxford repository Our World in Data (OWID(24, 25)) COVID-19 cases per population (black heatmap) were recapitulated by the CMU-CTIS self-reported recent positive COVID-19 tests per survey trends (navy heatmap, survey questions summarized in **Supplementary Table 1**) using Spearman correlations (left-most green annotation), excluding 2 countries without benchmark data. All countries in Europe were benchmarked whereas those in Africa, Asia and the Americas were benchmarked less frequently and with lower strength. It is presently not possible to distinguish lack of benchmark correlation due to poor UMD-CTIS performance versus lower quality benchmark data in a specific country. While survey versions can result in signal trend discontinuities, and should be kept in mind when utilizing UMD-CTIS outputs, this signal did not have notable discontinuities (also see additional benchmarks in **Supplementary Figure 1)**.

We then tested whether human social sensing — that is, using a proxy response of knowing someone with COVID-like-illness (CLI) in the respondents’ local community (community CLI, burgundy heatmap) — could amplify the COVID-19 case signal, thus providing meaningful trends in areas with lower survey coverage(21). More countries were benchmarked and with higher strength, even in lower-coverage countries where publication representation is also lower (e.g. Americas).

Lastly, we evaluated two syndromic surveillance signals, broad CLI (fever, cough or anosmia/ageusia in red heatmap) and narrow CLI (anosmia/ageusia with illness < 14 days duration in orange heatmap), that were strong and consistent test positivity predictors in prior analyses(26, 27). These signals performed similarly, with broad versus narrow stronger in some regions compared to others, providing potential signals for regions with strained testing capacity or utilization.

The peak week of each signal (scatter plot open shapes) was similar to the benchmark peak (scatter plot filled circle). Some signals peaked with a different wave in regions with multiple waves. Overall, UMD-CTIS signal trends reveal the global pandemic “fingerprint” from benchmark case trajectories, thus highlighting that limited questions and sampling can nevertheless enable regional public health tracking and insights.

### Predicting Global COVID-19 Impact Trends

We sought to determine whether we could build a model to predict test positivity utilizing symptoms (reported present in the prior day) and simple demographic categories, at the national and global scale. We hypothesized that a very large pooled global model would perform better than individual country-specific models, especially in regions with a paucity of testing data. After splitting the data, we used basic LightGBM to train and test country- and globe-specific models. We hyperparameter tuned the global model with grid search (**Supplementary Table 2**). We additionally tested country-specific models versus the global and vice versa. Using F1, the harmonic mean of precision and recall, the final global model on a balanced dataset performed well at 0.74 overall (0.71 test positive, 0.78 test negative, 0.7% improvement with tuning). Accuracy (0.82), precision (0.73), recall (0.75) and Area Under Curve (AUC, 0.799) are shown in the confusion matrix and Receiver Operating Characteristics curve (ROC) in **Figure 3**, and was comparable to that from a large prospective longitudinal data set(11). National models (F1 median [IQR] 0.71 [0.64, 0.75]) and global model tested on national data performed similarly (F1 median [IQR] 0.72 [0.66, 0.76]). Notably, the global model performed better in 77% countries and in 93% of below-median-F1-countries(**Figure 3**).

**Figure 3.**
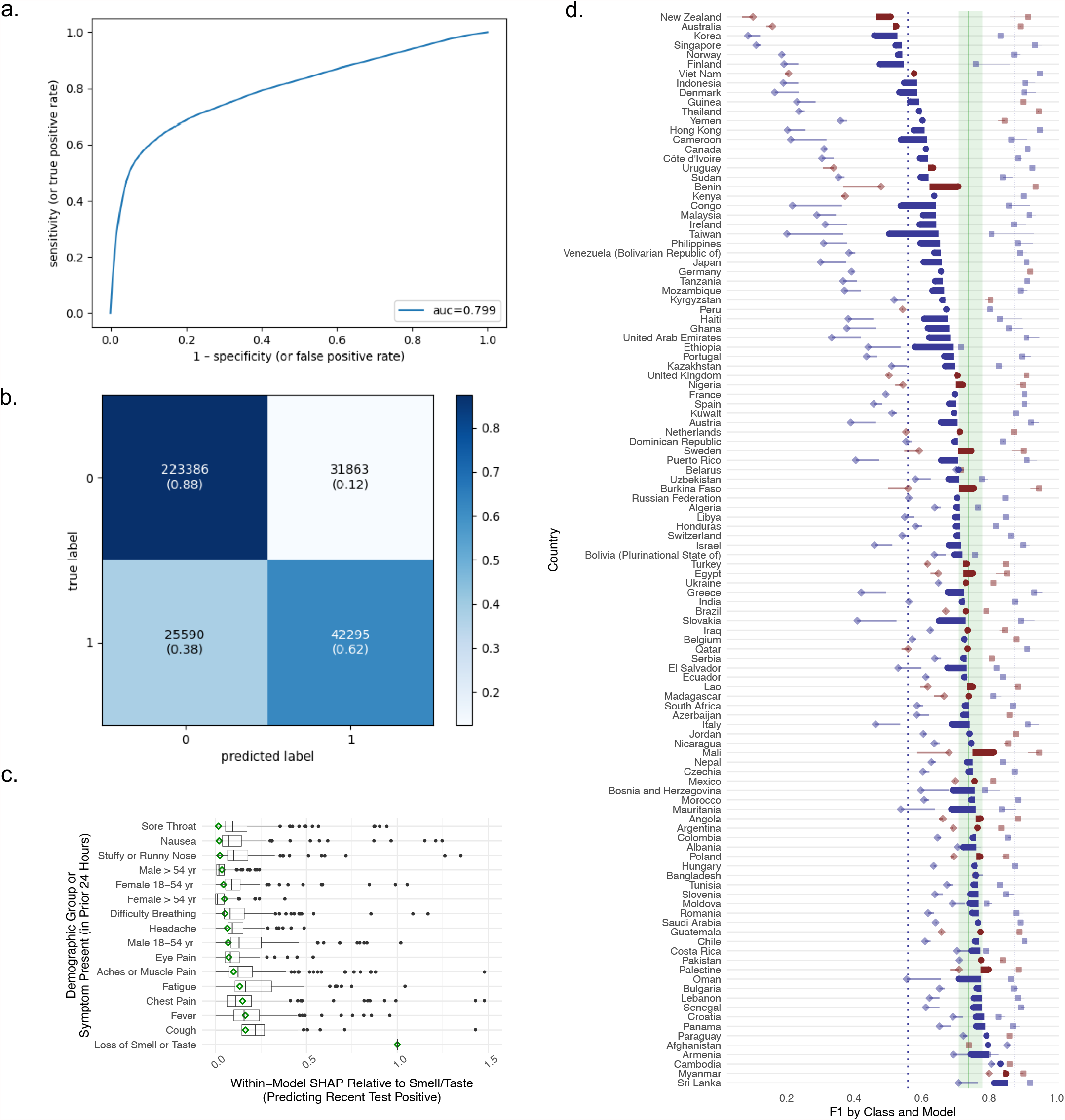
Model to predict recent COVID-19 test positivity using self-reported symptoms and minimal demographic data for the national and pooled global data. a) Area Under the Curve (AUC) for the Receiver Operating Characteristic (ROC) of the hyperparameter tuned global model. b) Confusion matrix for the hold-out global data. c) SHapley Additive exPlanations (SHAP) distribution of relative feature importance from the national models (box-and-whisker plots) compared to the global model (green diamonds). Within-model feature importance normalized to anosmia/ageusia to facilitate between-model comparison. d) Comparison of model performance using F1. Country-specific F1 overall (circle) and for negatives (class 0, diamond) or positives (class 1, square). Increase (blue) or decrease (red) in F1 with application of the global model to country data. F1 annotations shown for each class for median national (vertical blue dotted lines) and global (border of green shaded area) models, and overall global F1 (vertical green line).

We predicted test positive trends among all survey respondents, assuming a 0.5 probability threshold with the global model. This prediction model-based signal applied to survey responses without test data performed similarly to the test positivity signal (N=75 Spearman correlations > 0.4). The relative importance of input features, especially anosmia/ageusia followed by cough and fever, was consistent with prior studies(26, 27), and showed limited variation in 115 models (**Figure 3**). The young-male category was the strongest demographic feature, but other demographic features influenced many national models. Additionally, nausea, sore throat and stuff/runny nose were substantially less important in the global model than most regional models.

A similar approach for predicting test receipt was also conducted, though positive test prediction was much better than tested recently prediction (F1 0.57). This is likely due to spatiotemporal testing access and regulatory variability, and the omission of key predictors of testing receipt (e.g. contact, urban living) and was beyond the scope of this analysis.

### Risk-Mitigation Perception and Other COVID-19 Impact Trends

The cross-sectional FAUB sampling strategy in UMD-CTIS can inform public health officials on a range of impacts, such as community transmission, testing barriers, socio-economic insecurity, knowledge, practices and mitigation measures(18, 20, 28, 29). Despite the cross-sectional design, the order of symptom onset inferred from the distribution duration of symptoms among those with incident (< 14 duration) illness broadly was consistent with that from longitudinal studies, with loss of smell or taste last appearing later than most (**Supplementary Figure 3**)(30), and just after COVID-19 testing. These patterns, however, are incidence- and recall-dependent, and would not replace the quality of data from longitudinal studies, but could support efforts in resource limited areas, especially with the advent of a novel pathogen.

Here we also highlight the ability of the UMD-CTIS to show COVID-19 impact trends at a national scale, including regions that were under-represented by publications (**Figure 4**). In Nicaragua (enlarged panels), weekly trends in self-reported risk perception (i.e. community CLI or worry about COVID-19) and mitigation measures (i.e. no direct contact in prior day, masking in prior 7 days). In general, risk perception measures are positively correlated with each other and with protective measures. Some countries show an apparent lag in intensification or deintensification of protective measures following a rise or fall in being worried about COVID (grey panel horizontal axis, noting the non-zero intercept to show between-country trends), thus giving the appearance of a loop or spiral (e.g. Hungary), similar to other biological systems for which there is a delay before a stimulus can effect change or recovery. Social-distancing measures appear to have a stronger association (steeper curve) in some countries (e.g. El Salvador), suggesting the combination of social-distancing or masking (flatter curve) is more stable, suggesting there are population-level mitigation trade-offs(18, 31). In Tunisia, for example, this pattern was present early on, then shifted to a flatter association with social-distancing later, perhaps indicating that this mitigation measure, despite COVID-19 concern, may be less sustainable or mandated by regional governments. Lastly, while community CLI and worry about COVID-19 were largely positively correlated, this was not the case in all countries (e.g. Ethiopia), where worry and mitigation declined despite rising community CLI. These visualized relationships are undoubtedly driven by individual and population factors, and may not be causal. Given that mandates for social-distancing and masking are not uniformly effective, the ability to assess population-level concern could inform reopening and intensification of mitigation strategies. Also, the ability to appreciate country-specific patterns, lags or differences in self-perceived risk and behavior could be relevant when considering other mitigation strategies such as vaccination.

**Figure 4.**
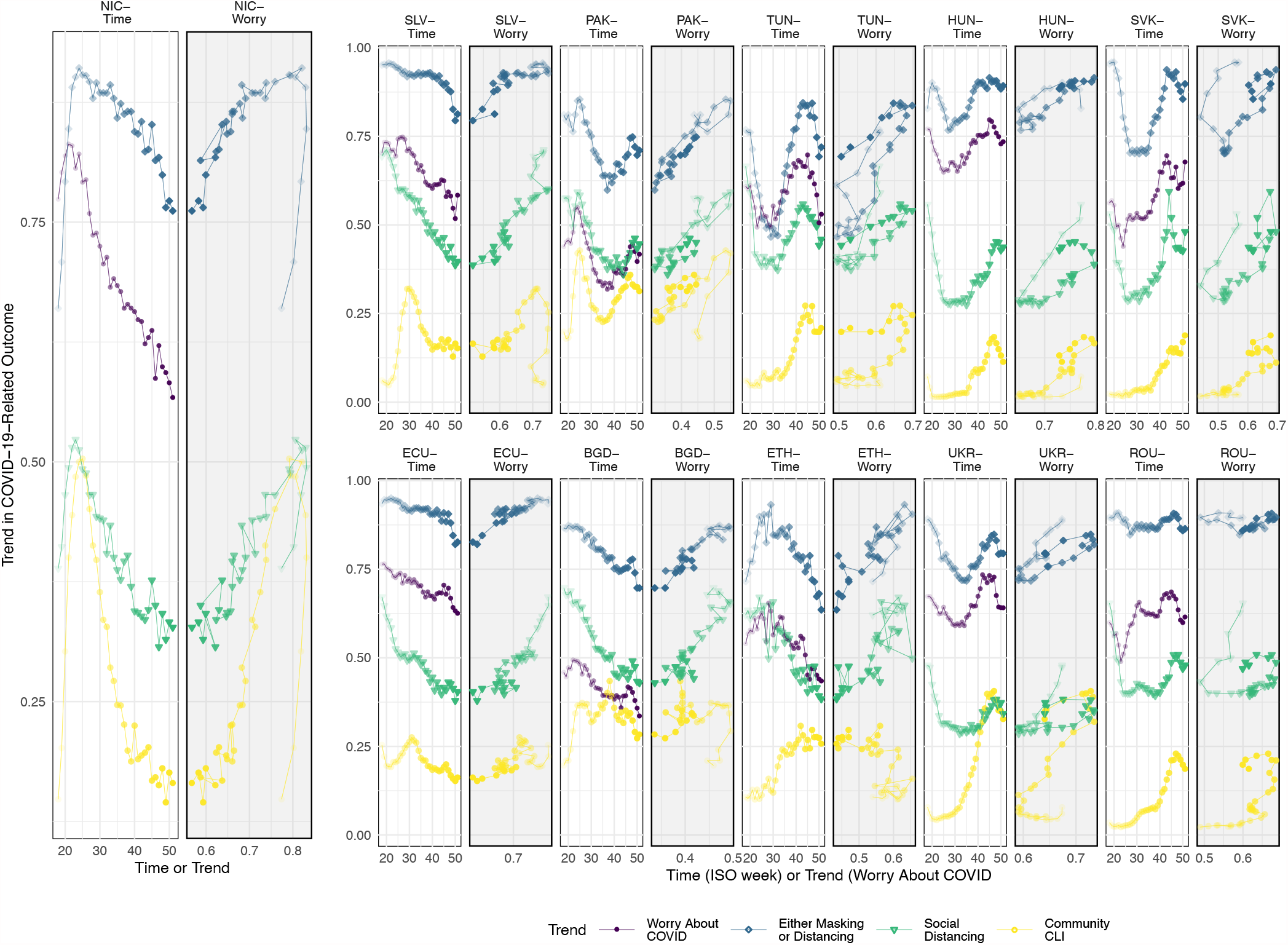
Illustration of the relationships between self-reported COVID-19 worry versus self-reported mitigation behaviors. For each example country (Nicaragua enlarged to show detail), the time series for four COVID-related impacts are shown on the left (white panel), while the relationship between worry about COVID-19 illness trends (horizontal axis) versus the other impacts (vertical axis) are shown on the right (grey panel). Worry about COVID-19 trends (purple circles) tends to be positively correlated with higher community CLI (knowing someone with CLI in the local community, yellow circle), an increase in social distancing (no direct contact in prior 24 hours, green triangles), and an increase in combined mitigation efforts (masking all or most of the time when out in public in the prior 7 days, not out in public in the prior 7 days, or social distancing, blue diamonds). For some countries (e.g. Hungary), a change in mitigation seems to lag behind a that in COVID-19 worry, creating a loop or spiral appearance (grey panel). In others, COVID-19 tracks with mitigation efforts, but seems to be unrelated to community CLI. Countries selected from those that benchmarked or were well-represented (see Figure 1), and to illustrate patterns. Each week is shown as a progressively darker shape from April 27 through December 20, 2020, by ISO week.

## Discussion

While many areas of the world have access to sophisticated, timely and reliable methods for tracking COVID-19 impacts over time, there are billions of people living in regions without these resources. And yet, a necessary step in dampening global COVID-19 transmission is the ability to efficiently and quickly monitor events and responses locally, across the globe, and especially as governments downsize disease surveillance programs(32). The UMD-CTIS online survey platform — delivered to social media users and leveraging Facebook-provided infrastructure and bolstered by public health partners and survey methodologists — has the potential to fill this need.

Here we show that cross-sectional responses to surveys offered daily to a statistical sample of over 2 billion Facebook social media users worldwide can provide COVID-19 impact trends that are correlated with benchmark COVID-19 case time series. We used limited survey questions and a pooled global dataset to benchmark and predict case trends, indicating that parsimonious surveys delivered with this novel sampling strategy, despite potential biases and country-specific heterogeneity, can provide meaningful trends and regional insights. Consistent with recent research(26) from this and two other platforms(10, 11, 33) in countries well-represented in the COVID-19 biomedical literature, self-reported loss of smell or taste, cough and fever in the prior day remain important signals and predictors of self-reported test positivity across a range of languages, cultures, COVID-19 waves and demographic differences using a unified survey platform. Individual country differences may be more pronounced with other types of analyses, and the point estimates may not be transferable to burden estimates(34), however, thus the value of this sampling scheme is with respect to the within-country trends and regional impacts. The time-series trends nevertheless recapitulated COVID-19 waves across the globe (**Figure 2**), and are proof of principle that UMD-CTIS could complement other data streams.

Importantly, the COVID-19 impact signal that was most often and most highly correlated with benchmark data was the answer to a single question: “Do you personally know anyone in your local community who is sick with a fever and either a cough or difficulty breathing?”. This simple signal, created by human social sensors(21), performs in regions with lower survey coverage, effectively scaling up the N through respondents’ CLI reporting on behalf of the local community. This proof of concept has been demonstrated in other settings(35). In contrast, more specific signals (e.g. loss of taste or smell, positive test), may be more relevant at higher COVID-19 incidence, when transmission is falling, in highly vaccinated populations, or with co-circulation. The UMD-CTIS survey could be deployed more intensively in areas that did not benchmark or in subregions requiring closer monitoring (e.g. poor vaccine coverage and routine government reporting stopped) with just this one community CLI question, and then supplemented with other survey questions at a coarser spatiotemporal scale.

Designed as a cross-sectional survey, UMD-CTIS cannot link repeated individual responses to evaluate individual level effects or disease trajectories. Because of the relative consistency of the sampling scheme, from a study base population, however, we are able show that average behavior of individual-level disease trajectories can be inferred, though not with the quality and precision of other platforms specifically design for this. Ecological bias must be considered, though in this study, the outcome of interest is primarily population-level trends, not individual-level causal effects. Because of the extensiveness of the FAUB and the sampling strategy, and the survey-weights, the cross-sectional UMD-CTIS is a robust representation of the population over time and may therefore be more generalizable than other opt-in surveillance methods or observational studies (e.g. hospital-based cohorts, longitudinal tracking application) for certain measures.

Syndromic surveillance in a pandemic may also have confounding, selection and measurement bias since generally health-engaged subjects participate and continue to participate over time, data are self-reported, digital interface access is necessary, and outbreaks may skew epidemiological parameter estimates(23, 36, 37). However, the powerful and efficient strategy of user-base sampling of a defined cohort and survey-weights to improve representativeness is unique among existing syndromic surveillance tools. Even in areas with less representation in the COVID-19 literature, many people utilize social media and may therefore be more accessible than traditional surveys.

Additional important considerations are the relatively high baseline of many metrics, perhaps due to response patterns, or so-called “trolling” behavior. We attempted to benchmark with minimal data manipulations, as these biases may be less likely to impact time trends, and the strength of this survey strategy is in time trends and the ability to conduct small area estimation (19). However, there can be trend breaks with survey versions (e.g. testing queried of symptomatic initially, then of all respondents), though these can be mitigated with sensitivity analyses (e.g. stratified by version), non-parametric correlations methods of trends rather than use of the specific point estimates. Other outputs, such as estimation of absolute burden or *Rt* would require methods to deconvolute cross-sectional responses and adjust for the baseline response rate.

Despite acknowledged possible limitations, the potential value of the UMD-CTIS is substantial. Compared to the Gallup World Poll, the largest global survey using established methodology, the UMD-CTIS survey is conducted on a daily (not annual) basis, and using a social-media based instrument instead of telephone and in-person interviews(38). Gallup World Poll interviews about 1000 subjects per country per year, while the UMD-CTIS sampled five to ten times that per country per week with a fraction of the in-country infrastructure and human resources(39). The great majority of the cost and logistical resources of the survey and deployment are borne by Facebook and the UMD Joint Program in Survey Methodology (JPSM). The purpose of the UMD-CTIS is not to provide exact and unbiased on-time point estimates, but to allow for the analysis of time trends and small area estimation. With this, UMD-CTIS is the largest ongoing real-time, remote, global health survey ever conducted. Importantly, the survey serves many countries that have been relatively under-represented in COVID-19 biomedical literature to date.

The other covariates in UMD-CTIS provide a valuable window into impacts on populations that may be underrepresented in the COVID-19 biomedical literature. We highlight in this study only a handful of these, including population-level concern about COVID-19-related illness, in relation to temporal changes in self-reported mitigation strategies. Each country experienced COVID-19 differently and yet there were still qualitative regional similarities. Population-level social distancing was positively correlated with worry about COVID-19 illness, hinting that perhaps intrinsic motivators are generalizable drivers of this behavioral modification. We do see features to suggest that mitigation behavior may lag risk perception, and that mitigation behavior choices may shift, similar to recovery from a stressor or vigilance fatigue.

As a digital health tool translated into dozens of languages for hundreds of countries, the UMD-CTIS survey has the ability to adapt to on-demand public health needs. With each survey version, new questions have been added in conjunction with epidemiologists, local government input and research partners. Other studies have used these data to investigate a range of insights from testing practices or vaccine hesitancy, and there is potential to further understand knowledge, attitudes and practices through this high-density data stream(28, 40–43). Indeed, the survey has most recently incorporated items to survey social impacts, vaccine uptake and vaccine hesitancy(29). With the expected return of seasonal respiratory infections(44) and the possibility that influenza burden may be doubly severe(45), as well as possible emergence of other known or novel pathogens of significance, the sampling strategy for this global health tool may be invaluable in conjunction with government reporting, viral surveillance and regional-specific syndromic surveillance methods(18, 20), if available. In conclusion, UMD-CTIS has shown to be a valid and powerful means of monitoring waxing and waning impacts of COVID-19, in a range of settings that include in under-represented countries. The publicly available aggregate as well as research-specific micro-data(33) are unique resources to track and enhance a range of public health efforts to control COVID-19 globally.

## Materials and Methods

### UMD-CTIS Survey

This research is based on survey results from UMD-CTIS, approved by the UMD Institutional Review Board (IRB, 1587016-10) and described previously(19). Briefly, each day, a subset of the 2.6 billion-person global Facebook active user base (FAUB) were sampled with replacement, and invited to participate via a special banner on the top of their news feed (summarized in Figure 1a). Sampled users who did not respond were given a resting period before resampling. Resampling of users who previously responded was theoretically possible, but quantitative resampling measures cannot be provided. After obtaining consent in Facebook, users were pushed to the Qualtrics platform utilizing a token system. All respondents must self-report age ≥ 18 years to participate. All demographics and survey items were collected by Qualtrics using a web-based, cross-sectional survey translated into commonly spoken languages in the N=250 countries surveyed. Qualtrics implemented standard survey administration methods (e.g. response randomization). Facebook provided survey weights that account for non-response and standardize for country-level demographics (N=114 countries). The survey was updated periodically with survey methodologist and epidemiologist input to address timely public health questions (e.g. masking behaviors, vaccine hesitancy), with several days overlap between each survey version. UMD compiled daily, real-time micro-data as well as publicly available survey-weighted aggregated data(33). Users of the micro-data can utilize the self-reported demographics to improve weighting adjustments.

### Study Design and Benchmarking

Retrospective cross-sectional analyses presented herein include micro-data survey responses from countries with survey-weights, from the first full International Organization for Standardization (ISO) week from UMD-CTIS inception (April 23, 2020). This study uses data from complete ISO weeks through version 6 (i.e. April 27, 2020 -December 20, 2020), and were accessed January 21, 2021. Surveys for versions 1-6 are available online(33), with key inputs used here summarized in Supplementary Table 1. This study was approved by the BCH IRB (P00023700). The number of UMD-CTIS surveys per capita were compared to COVID-19-related biomedical citations per capita, using the search logic: (“COVID-19” OR “SARS-CoV-2 ” OR “2019-nCoV” OR “ncov2019”) AND (Country Name OR Alternative Country Name). UMD-CTIS trends over time were compared to benchmark data from the publicly available OWID(24) accessed April 23, 2021. Survey-weighted proportions were calculated by country and day(46). Seven-day smoothing was applied to the time series. Survey questions without responses were coded as “not present”, “no” or “false”, except where noted (though some survey versions differentiate missing as seen but not answered versus not seen and not answered). Spearman correlations of OWID benchmark data versus UMD-CTIS signals were calculated.

The primary comparison was benchmark cases versus survey-weight test positives per survey (i.e. recent test resulted positive), community CLI (i.e. knowing CLI in respondent’s local community), broad CLI (i.e. fever, cough or anosmia/ageusia in prior 24 hours), narrow CLI (anosmia/ageusia and illness duration of < 14 days) and predicted test positive. Secondary comparisons included test positive proportion and tests. For trend visualization, smoothed country-day trends (Xc,t) were normalized to range from [0,1] using the [minimum, maximum] over the study period (T) for each country (i.e. Yc,t = (Xc,t -min(Xc,t over all T)/max(Xc,t -min(Xc,t over all T) over all T))), and were grouped into four geographic regions. Clustering for visualization used benchmark data (using row clustering method=”complete” in Heatmap from ComplexHeatmap 2.3.4, R 3.6.3 https://www.R-project.org/). We evaluated several approaches for simple cleaning including complete demographic data (age, sex and/or geographic sub-region), survey response time, weight outliers, and survey response patterns. For most analyses (exception below), we limited cleaning to highlight the utility of raw signals regardless of clearing approaches.

### Prediction of Global COVID-19 Test Positivity Trends

To enable evaluation of COVID-19 case trends with minimal testing data, we conducted a retrospective diagnostic classification analysis by training a gradient boosting decision tree binary classifier. Four base modeling methods (Logistic Regression, Gaussian Naive Bayes, Support Vector Machine, and Light Gradient Boosting Machine) were compared before determining that Light Gradient Boosting Machine (LightGBM, lightgbm in Python 3.8 with Shapley values visualization package for predictor visualization) was the highest performing at a moderate time cost, and is presented throughout.

The input features were self-reported age-gender categories (using self-reported age 18-54 versus ≥ 55 years, gender male versus female) and responses to 12 symptoms queried in versions 1-6. A fifth other age-gender category was coded if either age was missing, or gender was not “male” or “female”. Symptoms without responses were assumed to be “no” if at least one symptom response was provided.

Each of five covariate-specific strata for each country was split 3:1 into a training:validation. Since the test negative and positive data were inherently imbalanced (median [IQR] negative/positive 4 [2, 10]), and to limit over-fitting, the training set was down-or up-sampled to ensure balanced classes before 10-fold stratified cross-validation. The primary COVID-19 outcome variable was the self-reported positive versus negative recent COVID-19 test result (missing responses excluded), while a secondary analysis considered self-reported recent COVID-19 testing (missing responses assumed “no”). Sensitivity analysis including all missing values; strata of illness duration, demographic, and geographic data; symptom count; unusual symptoms; and excluding version 1-4 (change of survey logic June 26, 2020) had little impact on model metrics so were not utilized in the main model. Models were trained on each country and a pooled global data set. We then tested the global model on each individual country and vice versa.

## Data Availability

Anonymous responses and survey weights are processed at the University of Maryland, which also oversees survey content. Aggregated data are released to the public in near-real-time. Researchers may apply to use raw micro-data to study COVID-19 through a Data Use Agreement (https://covidmap.umd.edu/fbsurvey/)

## Acknowledgments

Facebook Sponsored Research Agreement INB1116217 (CMA, JSB, ELC, BR, GT and KAM (Mc Cord -De Iaco)). Facebook, Inc. Grant No. 4332732 (SLC, XD, KS, FK). The authors would like to acknowledge the programmatic support of Kara Sewalk and Aimee Han for this research.

## Supplement

**Fig. S1.**
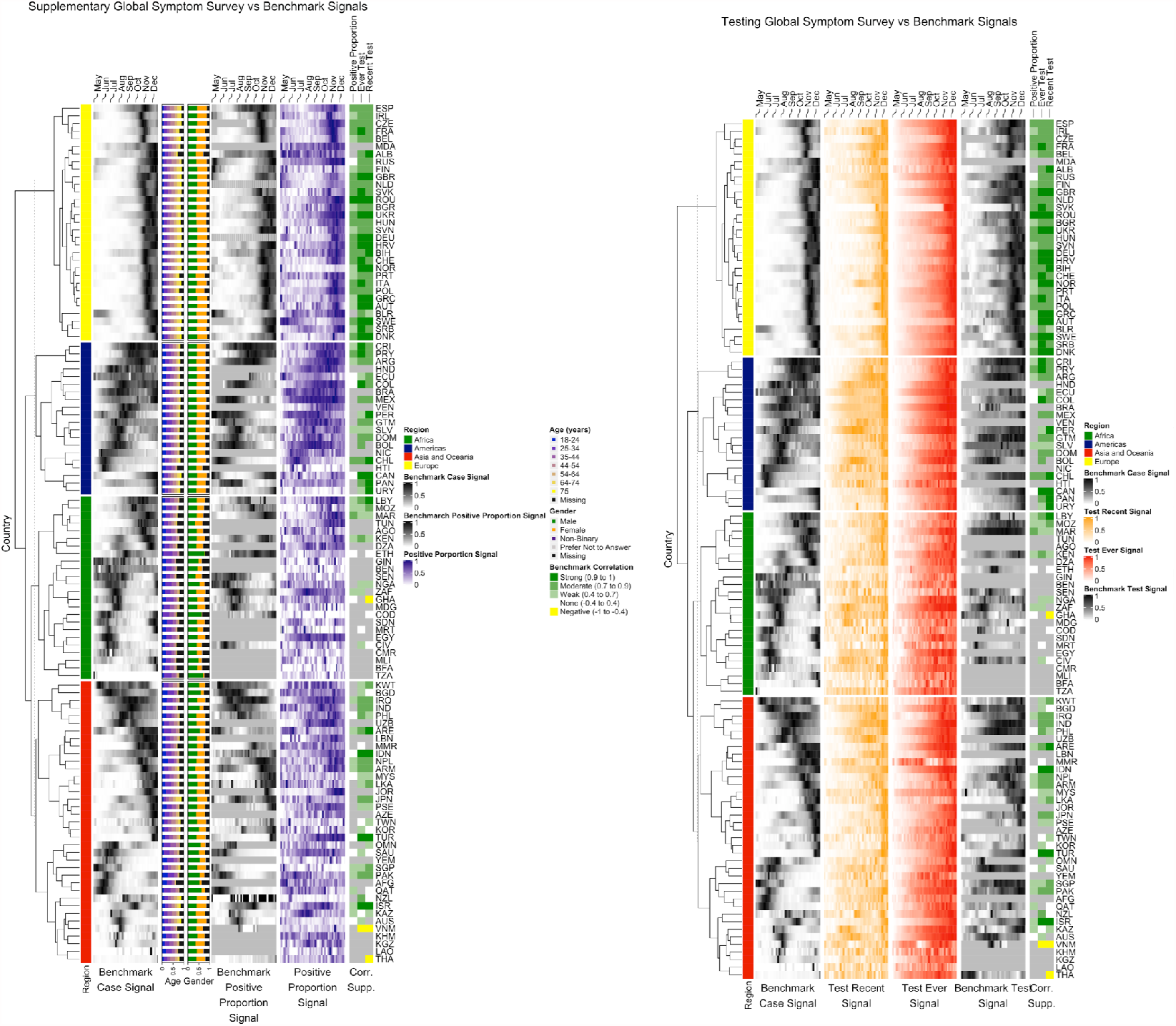
Heatmap of alternative signals and demographic data. Age and gender distribution after application of survey weights. Test recent (among the symptomatic), test ever and test positive proportion versus benchmark signals of cases, tests, and test positivity. All signals are normalized to range from 0 (min) to 1 (max) signal for between-country and temporal comparisons of trends. Benchmark correlations annotated using Spearman correlation. Note there was a change in testing queried after version 1-4, with a slight discontinuity with version change depending on which signal is evaluated. Discontinuities are possible with any version change, or even non-survey factors (e.g. phone software update) and must be taken into consideration when utilizing survey signal trends. Test recent was expanded to all respondents which created a trend break. Test recent was therefore restricted to the symptomatic for the purpose of signal comparison.

**Fig. S2.**
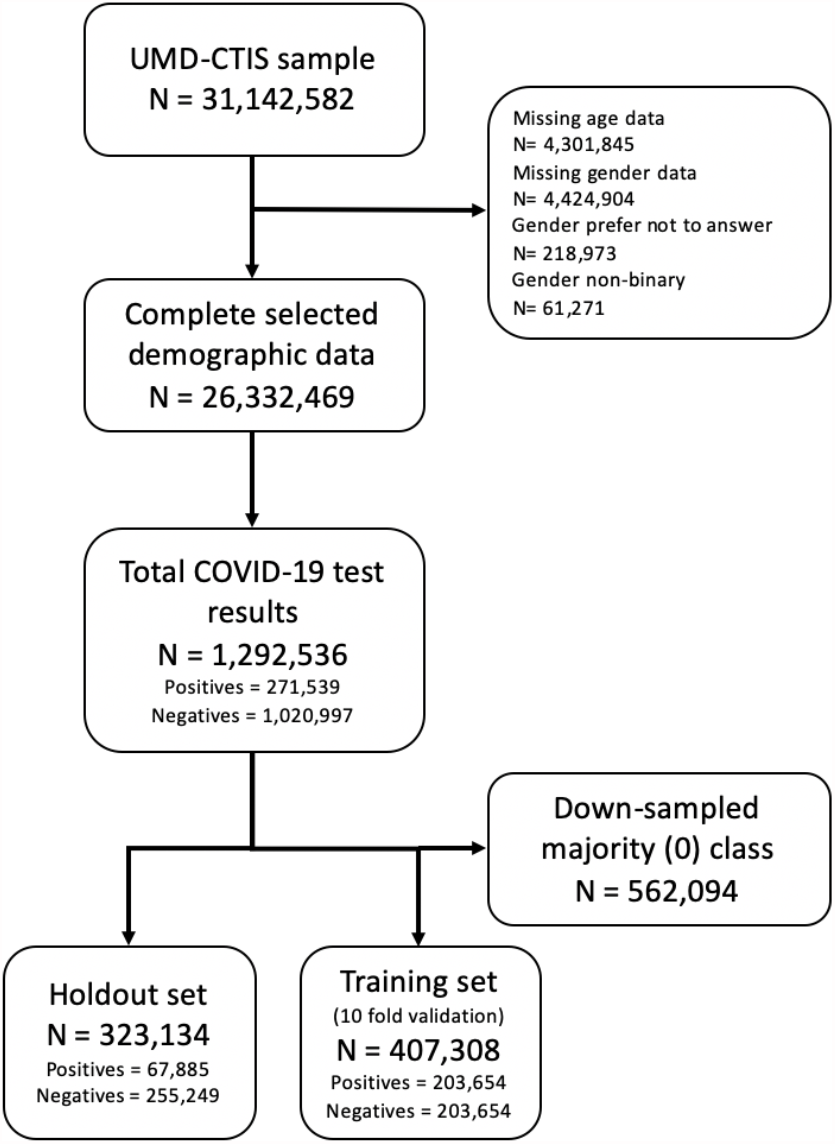
Flow diagram of the predictive model data set. Diagram showing the full sample, and subsampling required to obtain a labelled data set for machine learning using symptoms and basic demographic factors. Down-sampling of the majority class to limit over-fitting was conducted after splitting the data into testing and validation sets, within each demographic group.

**Figure S3.**
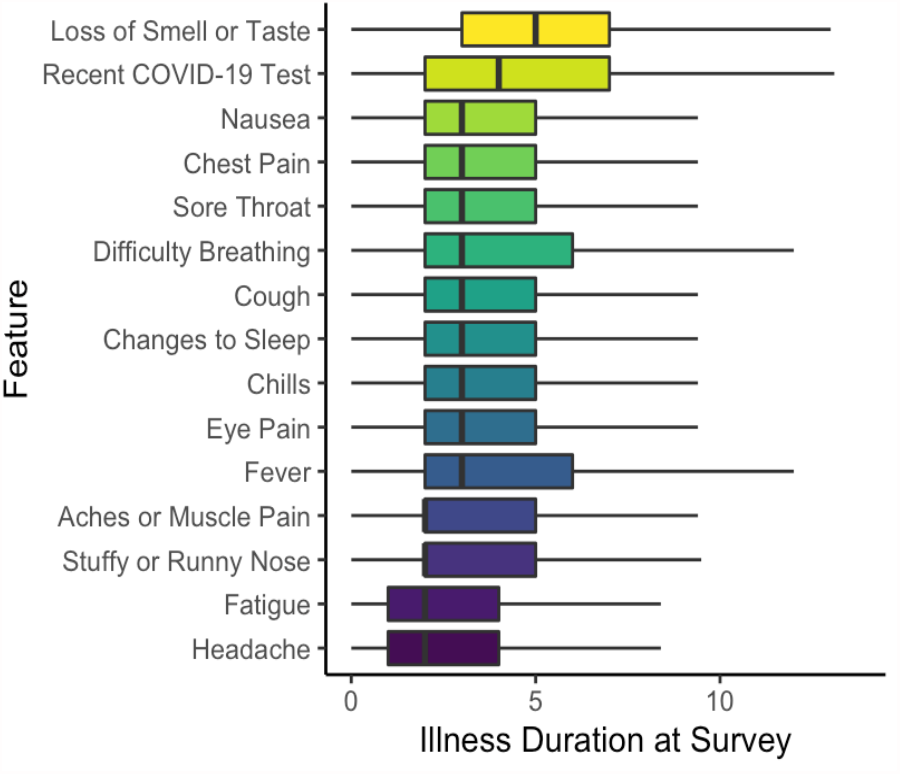
Oder of appearance of symptoms and reported testing among symptomatic respondents, for the pooled global sample during the study period, inferred from cross-sectional surveys and duration of illness at survey. Box-and-whisker plots of Inferred Order of Potential COVID-19 Proxies from UMD-CTIS. Among respondents with a self-reported incident illness (i.e. duration of illness of 14 days or fewer), inferred order of appearance of each feature using the distribution of illness duration from cross-sectional surveys (as symptoms were ascertained if present in the prior 24 hours during periods of COVID-19 transmission).

**Table S1.** Survey questions, response options, and notes regarding survey changes and coding.

| Covariate | Survey question/description | Response | Comments, versions and changes |
| --- | --- | --- | --- |
| Symptoms | B1: In the last 24 hours, have you had any of the following?<br>Fever, cough, difficulty breathing, fatigue, stuffy or runny nose, aches or muscle pain, sore throat, chest pain, nausea, loss of smell or taste, eye pain, headache. | Respondents may select none or up to 14 symptoms<br>1 = Yes<br>2 = No | Chills (version 3) and changes in sleep (version 6) were not included in our analyses since they were added later and this presented with many missing values. |
| Duration of illness | B2: For how many days have you had at least one of these symptoms? | Open response:<br>number validation | Only asked if B1 selected symptoms count greater than or equal to 1 |
| Community COVID-19-Like Illness (CCLI) | B3: Do you personally know anyone in your local community who is sick with a fever and at least one other symptom? | 1 = Yes<br>2 = No | In version 6 this was slightly changed to “Do you personally know anyone in your local community who is sick with a fever and either a cough or difficulty breathing?” |
| COVID-19 testing ever received | B6: Have you ever been tested for coronavirus (COVID-19)? | 1 = Yes<br>2 = No |  |
| COVID-19 testing recently received | B7: Have you been tested for coronavirus (COVID-19) in the last $\{e://Field/sympdays\}$ days?<br>Note: sympdays is B2, but capped at 14 days. | 1 = Yes<br>2 = No | Only asked if B6 is 'Yes' and B2 greater than or equal to 1 until version 4 (06/26/2020): then only asked if B6 is 'Yes' This was changed (version 7) to simply “Have you been tested for coronavirus (COVID-19) in the last 14 days?” |
| COVID-19 positive test | B8: Did your most recent test find that you had coronavirus (COVID-19)? | 1 = Yes<br>2 = No<br>3 = I don't know | Only asked if B7 is 'YES' |
| Social Distancing | C1_m: In the last 24 hours, have you had direct contact with anyone who is not staying with you? Direct contact means spending longer than one minute within two meters of someone or touching, including shaking hands, hugging, or kissing. | 1 = Yes<br>2 = No | Surveys answering NO coded as social distancing (per survey) |
| Either Social Distancing or Masking | C5: In the last 7 days, how often did you wear a mask when in public? | 1=All of the time<br>2=Most of the time<br>3=Some of the time<br>4=A little of the time<br>5=None of the time<br>6=I have not been in public during the last 7 days | Surveys answering 1, 2 or 6, OR those social distancing (see above) coded as either social distancing or masking (per survey) |
| Gender | E3: What is your gender? | 1 = Male<br>2 = Female<br>3 = Prefer to self-describe: [text entry]<br>4 = Prefer not to answer | This was changed in version 6 to<br>1=Male<br>2=Female<br>3=Other<br>4=Prefer not to answer |
| Age | E4: What is your age? | 1 = 18-24 years<br>2 = 25-34 years<br>3 = 35-44 years<br>4 = 45-54 years<br>5 = 55-64 years<br>6 = 65-74 years<br>7 = 75 years or older |  |
| Country | Country name as used in country level aggregates | Character | Ex: United Kingdom |
| Date recorded | Date that the response was recorded | Date/Time | Ex: 2020-05-01, 00:04:26 |
| Survey duration (time) | Duration of response in seconds | Integer | Ex: 72 |
| Weight | Survey weight to adjust to FB user population | Number (float) | Ex: 1526.36144<br>115 countries are weighted.<br>Weighting documentation:<br>https://arxiv.org/abs/2009.14675 |
| Missing data was coded throughout the survey as follows: -99 = missing/valid skipped/invalid,<br>-77 = seen but unanswered |  |  |  |

**Table S2.**
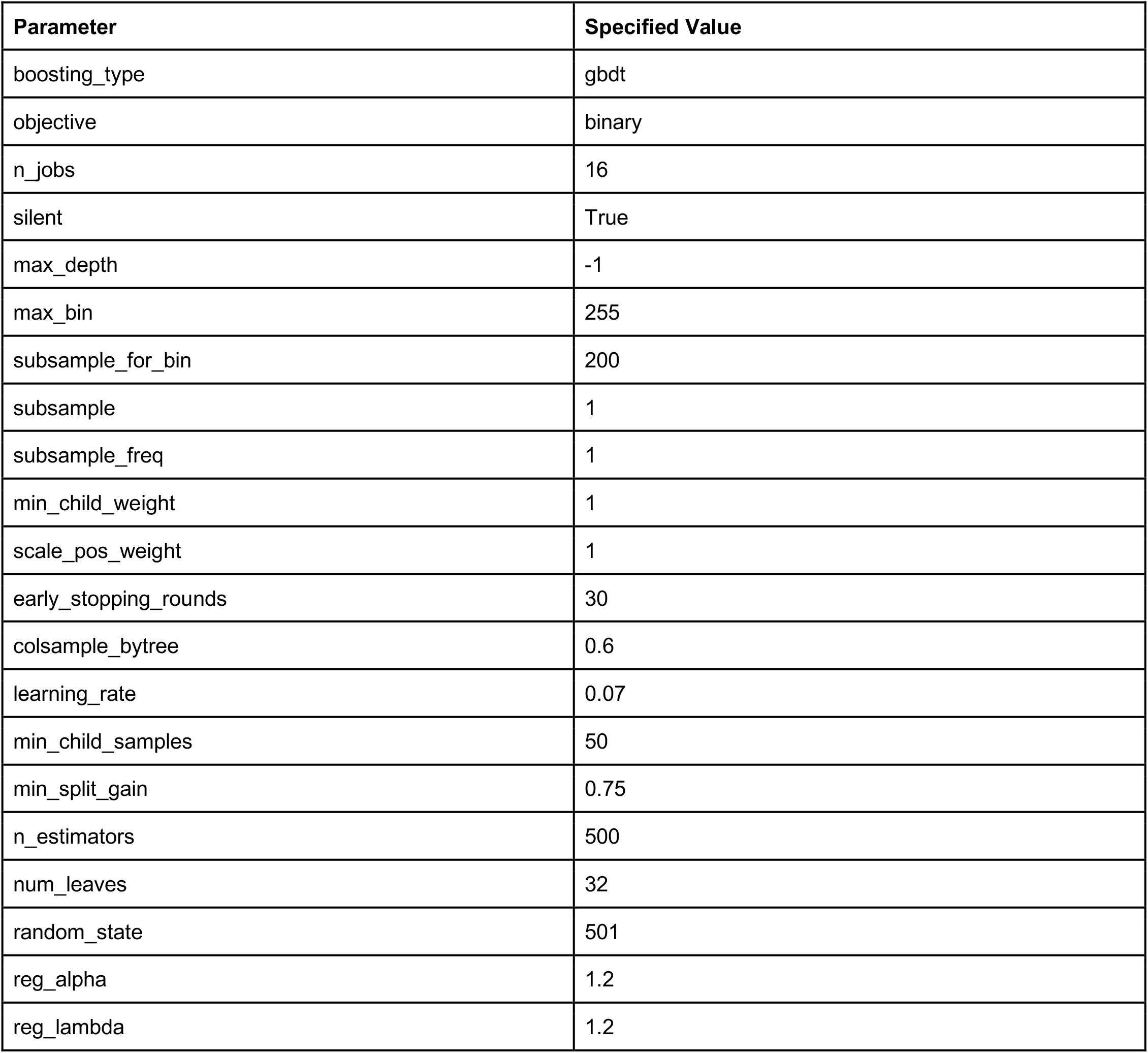
Parameters for global model. Specific *lgbm*.*LGBMClassifier* function parameters for the global model.

| Parameter | Specified Value |
| --- | --- |
| boosting_type | gbdt |
| objective | binary |
| n_jobs | 16 |
| silent | True |
| max_depth | -1 |
| max_bin | 255 |
| subsample_for_bin | 200 |
| subsample | 1 |
| subsample_freq | 1 |
| min_child_weight | 1 |
| scale_pos_weight | 1 |
| early_stopping_rounds | 30 |
| colsample_bytree | 0.6 |
| learning_rate | 0.07 |
| min_child_samples | 50 |
| min_split_gain | 0.75 |
| n_estimators | 500 |
| num_leaves | 32 |
| random_state | 501 |
| reg_alpha | 1.2 |
| reg_lambda | 1.2 |

## References

1. M. Lipsitch, D. L. Swerdlow, L. Finelli, Defining the Epidemiology of Covid-19 — Studies Needed. N. Engl. J. Med. 382, 1194–1196 (2020).

2. H. Tian, et al., An investigation of transmission control measures during the first 50 days of the COVID-19 epidemic in China. Science (80-.). 368, 638–642 (2020).

3. M. U. G. Kraemer, et al., Data curation during a pandemic and lessons learned from COVID-19. Nat. Comput. Sci. 1, 9–10 (2021).

4. N. A. Alwan, Surveillance is underestimating the burden of the COVID-19 pandemic. Lancet 396, e24 (2020).

5. E. J. Emanuel, et al., Fair Allocation of Scarce Medical Resources in the Time of Covid-19. N. Engl. J. Med. 382, 2049–2055 (2020).

6. J. Hellewell, et al., Feasibility of controlling COVID-19 outbreaks by isolation of cases and contacts. Lancet Glob. Heal. 8, e488–e496 (2020).

7. A. Güemes, et al., A syndromic surveillance tool to detect anomalous clusters of COVID-19 symptoms in the United States. Sci. Rep. 11 (2021).

8. L. Lapointe-Shaw, et al., Web and phone-based COVID-19 syndromic surveillance in Canada: A cross-sectional study. PLoS One 15, e0239886 (2020).

9. W. E. Allen, et al., Population-scale longitudinal mapping of COVID-19 symptoms, behaviour and testing. Nat. Hum. Behav. 4, 972–982 (2020).

10. H. Rossman, et al., A framework for identifying regional outbreak and spread of COVID-19 from one-minute population-wide surveys. Nat. Med. 26, 634–638 (2020).

11. C. Menni, et al., Real-time tracking of self-reported symptoms to predict potential COVID-19. Nat. Med. 26, 1037– 1040 (2020).

12. D. Yoneoka, et al., Large-scale epidemiological monitoring of the COVID-19 epidemic in Tokyo. Lancet Reg. Heal. - West. Pacific 3, 16 (2020).

13. S. J. Carlson, D. N. Durrheim, C. B. Dalton, Flutracking provides a measure of field influenza vaccine effectiveness, Australia, 2007-2009. Vaccine 28, 6809–6810 (2010).

14. M. S. Smolinski, et al., Flu near you: Crowdsourced symptom reporting spanning 2 influenza seasons. Am. J. Public Health 105, 2124–2130 (2015).

15. A. T. Chan, J. S. Brownstein, Putting the Public Back in Public Health — Surveying Symptoms of Covid-19. N. Engl. J. Med. 383, e45 (2020).

16. T. Varsavsky, et al., Detecting COVID-19 infection hotspots in England using large-scale self-reported data from a mobile application: a prospective, observational study. Lancet Public Heal. 6, e21–e29 (2021).

17. C. H. Sudre, et al., Attributes and predictors of long COVID. Nat. Med. 27, 626–631 (2021).

18. B. Rader, et al., Mask-wearing and control of SARS-CoV-2 transmission in the USA: a cross-sectional study. Lancet Digit. Heal. 3, e148–e157 (2021).

19. F. Kreuter, Partnering withturn-aroundfacebook globalon a university-basedsurvey rapid. Surv. Res. Methods 14, 159–163 (2020).

20. M. S. Graham, et al., Knowledge barriers in the symptomatic-COVID-19 testing programme in the UK: an observational study. medRxiv, 2021.03.16.21253719 (2021).

21. M. Galesic, et al., Human social sensing is an untapped resource for computational social science. Nature, 1–9 (2021).

22. K. Korkeila, et al., Non-response and related factors in a nation-wide health survey. Eur. J. Epidemiol. 17, 991–999 (2001).

23. K. Baltrusaitis, et al., Determinants of Participants’ Follow-Up and Characterization of Representativeness in Flu Near You, A Participatory Disease Surveillance System https://doi.org/10.2196/publichealth.7304 (July 4, 2021).

24. Oxford, Our World In Data. https://ourworldindata.org/ (April 23, 2021).

25. J. Hasell, et al., A cross-country database of COVID-19 testing. Sci. Data 7, 1–7 (2020).

26. C. H. Sudre, et al., Anosmia and other SARS-CoV-2 positive test-associated symptoms, across three national, digital surveillance platforms as the COVID-19 pandemic and response unfolded: An observation study. medRxiv, 2020.12.15.20248096 (2020).

27. D. Pierron, et al., Smell and taste changes are early indicators of the COVID-19 pandemic and political decision effectiveness. Nat. Commun. 11, 1–8 (2020).

28. J. Lessler, et al., Household COVID-19 risk and in-person schooling. Science (80-.). 372, 1092–1097 (2021).

29. D. Urrunaga-Pastor, et al., Cross-sectional analysis of COVID-19 vaccine intention, perceptions and hesitancy across Latin America and the Caribbean. Travel Med. Infect. Dis. 41, 102059 (2021).

30. G. Spinato, et al., Alterations in Smell or Taste in Mildly Symptomatic Outpatients with SARS-CoV-2 Infection. JAMA - J. Am. Med. Assoc. 323, 2089–2091 (2020).

31. S. Flaxman, et al., Estimating the effects of non-pharmaceutical interventions on COVID-19 in Europe. Nature 584, 257–261 (2020).

32. D. Fisher, A. Wilder-Smith, The global community needs to swiftly ramp up the response to contain COVID-19. Lancet 395, 1109–1110 (2020).

33. U. of Maryland, Repository of the COVID-19 World Symptoms Survey. https://covidmap.umd.edu/fbsurvey/ (mJanuary 21, 2021).

34. V. C. Bradley, et al., Are We There Yet? Big Data Significantly Overestimates COVID-19 Vaccination in the US (2021) (July 4, 2021).

35. C. Baquero, et al., The CoronaSurveys System for COVID-19 Incidence Data Collection and Processing. Front. Comput. Sci. 3, 52 (2021).

36. M. Lipsitch, et al., Potential biases in estimating absolute and relative case-fatality risks during outbreaks. PLoS Negl. Trop. Dis. 9, 1–16 (2015).

37. G. J. Griffith, et al., Collider bias undermines our understanding of COVID-19 disease risk and severity. Nat. Commun. 11 (2020).

38. Gallup, How Does the Gallup World Poll Work? https://www.gallup.com/178667/gallup-world-poll-work.aspx (2021) (July 3, 2021).

39. Gallup, Country Data Set Details. https://www.gallup.com/services/177797/country-data-set-details.aspx (July 3, 2021).

40. E. Gakidou, et al., Global projections of lives saved from COVID-19 with universal mask use. medRxiv, 1–35 (2020).

41. J. G. Lu, P. Jin, A. S. English, Collectivism predicts mask use during COVID-19. Proc. Natl. Acad. Sci. U. S. A. 118(2021).

42. G. Leech, et al., Mass mask-wearing notably reduces COVID-19 transmission. medRxiv, 2021.06.16.21258817 (2021).

43. E. Badillo-Goicoechea, et al., Global Trends and Predictors of Face Mask Usage During the COVID-19 Pandemic (2020) (July 3, 2021).

44. A. S. Maharaj, et al., The effect of seasonal respiratory virus transmission on syndromic surveillance for COVID-19 in Ontario, Canada. Lancet Infect. Dis. 21, 593–594 (2021).

45. B. D. Singer, COVID-19 and the next influenza season. Sci. Adv. 6, 86–115 (2020).

46. N. Barkay, et al., Weights and Methodology Brief for the COVID-19 Symptom Survey by University of Maryland and Carnegie Mellon University, in Partnership with Facebook (2020) (July 3, 2021).

